# Carrier frequency estimation of pathogenic variants of autosomal recessive and X-linked recessive Mendelian disorders using exome sequencing data in 1,642 Thais

**DOI:** 10.1101/2023.06.12.23291300

**Authors:** Wanna Chetruengchai, Prasit Phowthongkum, Vorasuk Shotelersuk

**Affiliations:** Excellence Center for Genomics and Precision Medicine, King Chulalongkorn Memorial Hospital, the Thai Red Cross Society, Bangkok, 10330, Thailand; Center of Excellence for Medical Genomics, Medical Genomics Cluster, Department of Pediatrics, Faculty of Medicine, Chulalongkorn University, Bangkok, 10330, Thailand; Division of Medical Genetics and Genomics, Department of Medicine, Faculty of Medicine, Chulalongkorn University, Bangkok, Thailand

## Abstract

People with autosomal recessive disorders often were born without awareness of the carrier status of their parents. The American College of Medical Genetics and Genomics (ACMG) recommends screening 113 genes known to cause autosomal recessive and X-linked conditions in couples seeking to learn about their risk of having children with these disorders to have an appropriate reproductive plan. Here, we analyzed the exome sequencing data of 1,642 unrelated Thai individuals to identify the pathogenic variant (PV) frequencies in genes recommended by ACMG. The ascertainment bias was controlled by excluding the carriers of the PV in the genes for the conditions that are attributed to their offspring disorders. In the 113 ACMG-recommended genes, 165 PV and likely PVs in 60 genes of 559 exomes (34%, 559/1642) were identified. The carrier rate was increased to 39% when glucose-6-phosphate dehydrogenase (G6PD) was added. The carrier rate was still as high as 14.7% when thalassemia and hemoglobinopathies were excluded. In addition to thalassemia, hemoglobinopathies, and G6PD deficiency, carrier frequencies of >1% were found for Gaucher disease, primary hyperoxaluria, Pendred syndrome, and Wilson disease. Nearly 2% of the couples were at risk of having offsprings with the tested autosomal recessive conditions. The expanded carrier screening focused on common autosomal recessive conditions in Thai seems to be benefit among the study samples.

## Introduction

Although each Mendelian disease is generally uncommon, with >5,000 known heritable disorders, they are accounted for ∼20% of infant mortality, ∼18% of pediatric hospitalizations, and substantial numbers in adults [1]. At least 2,000 autosomal recessive diseases [2] and 500 X-linked diseases [3] have been identified. The preconception carrier screening for these disorders and proper reproductive option counseling have been shown to be cost-effective measures that reduce the burden of Mendelian diseases [4-7]. The studies of the carrier rate of these disorders are highly varied with the number of tested genes and the genes included in the studies among different populations with no consensus agreement [8-11]. Until recently, the American College of Medical Genetics and Genomics (ACMG) released a set of 113 genes, both autosomal recessive and X-linked conditions, proposed as a standard list of genes for carrier screening [12]. Because X-linked glucose-6-phosphate dehydrogenase (G6PD) deficiency is the most common genetic disorder in Thailand and Southeast Asia [13], we added G6PD into the list and determined the carrier frequency and variant distribution of 114 recessive genes in the Thai population using the exomes of 1,642 unrelated Thais.

## Results

Using 1,642 exome sequencing data of unaffected parents whose children had rare diseases, we first excluded pathogenic and likely pathogenic variants (PV and LPV, respectively) found in these exomes, which caused the diseases in their children. We excluded 13 PV and LPV in 18 exomes (1%, 18/1642) (S1 Table).

Of the 113 ACMG-recommended genes, 165 PV and LPV in 60 genes of 559 exomes were found (559/1642: 34%). For G6PD, 7 PV and LPV of 127 exomes (female) were identified. In the analysis of a total of 114 genes, 172 PV and LPV in 61 genes of 640 exomes (640/1642: 39%) were identified (Table 1). Although we excluded HBA1, HBA2, and HBB genes causing thalassemia and hemoglobinopathies, the carrier rate of at least one screened disorder was still as high as 14.7% (241/1642) in our cohort. Of the samples, 7.4% (121/1642) carried more than one PV and LPV (Fig 1 and S2 Table).

**Fig 1.**
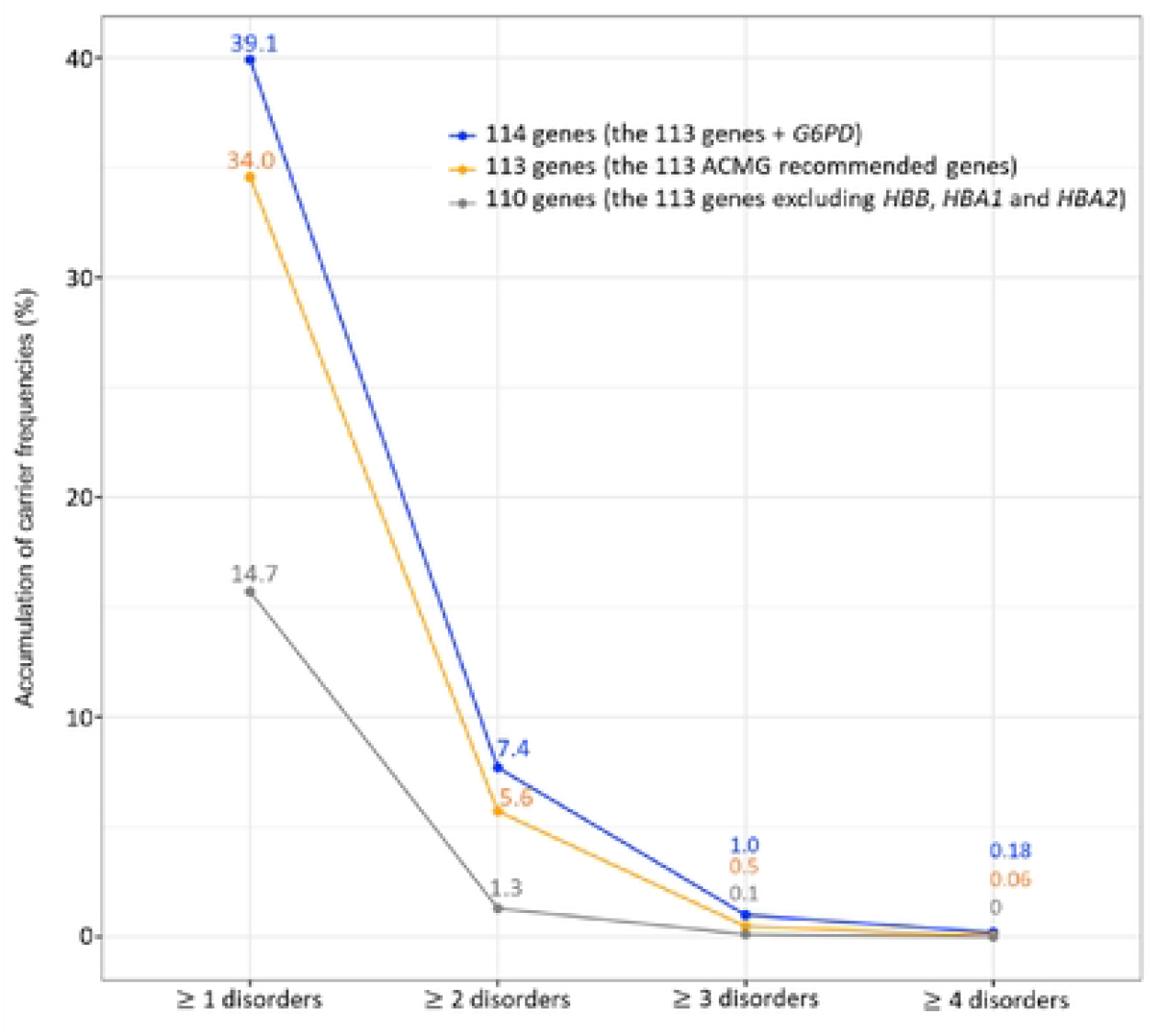
Carrier frequencies of one or more genetic disorders in 114 genes of 1,642 unrelated Thais. Orange color represents carrier frequencies in 113 ACMG recommended genes. Blue color represents those in 114 genes, which are the 113 genes and G6PD. Gray color represents those in 110 genes, which are the 113 genes excluding three genes underlying thalassemia and hemoglobinopathies (*HBB, HBAJ*, and *HBA2*).

### Gene carrier rate

In our cohort, all top three gene carrier rates were in hemolytic disorders with positive selection from malaria. The gene with the highest carrier rate is HBB, with 50.2% (321/640) and 19.6% (291/1642) of all carriers and participants, respectively. The second most common carrier gene is G6PD, with 19.8% (127/640) and 7.7% (127/1642) of all carriers and participants, respectively. HBA2 is the third most common gene, with 10.2% (65/640) and 4% (65/1642) of all carriers and participants, respectively.

There were 17 autosomal recessive genes with carrier frequencies of at least 1/250, an expected prevalence at birth of at least 1 in 250,000. As shown in Fig 2, three, five, and nine of these genes are related to hemolytic disorders (G6PD, HBB, and HBA2), inborn errors of metabolism (AGXT, ATP7B, CYP27A1, GBA, and PAH), and in the miscellaneous group (CEP290, CFTR, CLCN1, GJB2, MCPH1, NEB, PKHD1, SLC26A4, and USH2A), respectively.

**Fig 2.**
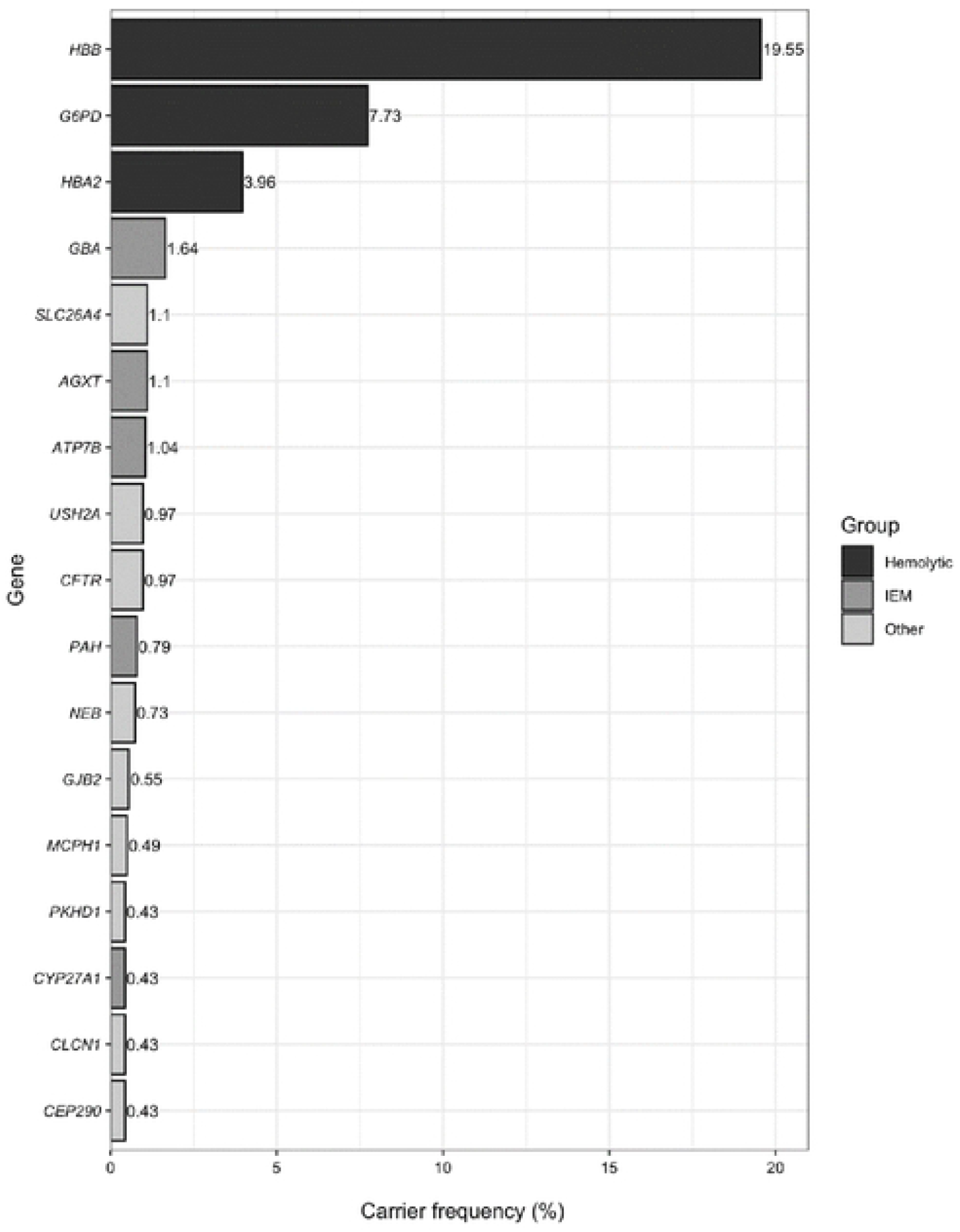
The 17 genes underlying autosomal recessive disorders with carrier frequencies of ≥ 1/250 (0.4%) in the 1,642 unrelated Thai individuals.

### Variant carrier rate

The most common PV is HBB; c.79G>A (p.Glu27Lys) responsible for hemoglobin (Hb) E (HbE), representing 45.5% (291/640) and 17.7% (291/1642) of all carriers and participants, respectively. HBA2; c.427T>C (p. Ter143Glnext*31, also known as Hb Constant Spring) is the second most common variant, representing 9.2% (59/640) and 3.6% (59/1642) of all carriers and participants, respectively. The third most common variant is G6PD; c.961G>A (p.Val321Met) causing G6PD deficiency, with 8.6% (55/640) and 3.4% (55/1642) of all carriers and participants, respectively. The variant carrier rates are shown in S3 Table.

### Thalassemia and hemoglobinopathies

Although, the most common type of α-thalassemia mutation is the deletion of one or more of the α-globin genes, HBA1 and HBA2 [14], we did not attempt to analyze this type of genetic variant due to the inherent limitation of exome sequencing. The carrier rates of nondeletional α-thalassemia were detected in 5.1% (71/1642; 6 in HBA1 and 65 in HBA2) of our cohort. The most common variant is c.427T>C 3.6% (59/1642) in Hb Constant Spring, followed by Hb Paksé c.429A>T 0.3% (5/1642). The frequency rate of β-thalassemia carriers is 19.6% (321/1642) with c.79G>A (p.Glu27Lys [HbE]) as the most common variant (17.7%, 291/1642), and HBB c.52A>T (0.79%, 13/1642c.126_129del (0.67%, 11/1642) were the next most common variant.

### G6PD deficiency

The carrier rate of G6PD variants was 7.7% (127/1642). The prevalence of heterozygous/carrier G6PD variant and homozygous females in our cohort is 15.28% (127/831) and 0.8% (7/831), respectively (S5 Table). The percentage of symptomatic or biochemical G6PD deficiency in these samples was not known. The prevalence of hemizygous G6PD variant males in our cohort is 8% (65/811). The female carrier rate corresponds with the predicted prevalence calculated with male hemizygous prevalence using the Hardy–Weinberg formula. c.961G>A (p.Val321Met) (G6PD Viantien) and c.577G>A (p.Gly193Ser) (G6PD Mahidol) are the two most common G6PD variants found in our cohort.

### Other autosomal recessive disorder carrier rate and common variants

The carrier frequency of PV and LPV in GBA causing Gaucher disease is 1.6% (27/1642) with c.605G>A (p.Arg202Gln) (0.54%, 9/1642) and c.1448T>C (p.Leu483Pro) (0.42%, 7/1642) being the two most common variants.

The carrier frequency of PV and LPV in genes associated with hereditary syndromic hearing loss SLC26A2 is 1.1% (18/1642). c.919-2A>G was the most common variant (0.3%, 5/1642). The second most common gene causing syndromic hearing loss carriers was USH2A (Usher syndrome) (0.97%, 16/1642). The carrier frequency of PV and PLV in GJB2, one of the most common genes causing nonsyndromic deafness is 0.6% (9/1642). All cases identified carried c.235delC (p.Leu79CysfsTer3) in GJB2.

Besides Gaucher disease, the most common inherited metabolic disease gene carriers in our cohort are hyperoxaluria (AGXT; 1.1%, 18/1642), Wilson disease (ATP7B; 1%, 17/1642), and phenylketonuria (PAH; 0.79%, 13/1642).

## Discussion

Next Generation Sequencing (NGS) has been increasingly used for expanded genetic carrier screening for multiple genes in a highly efficient manner in clinical laboratories [15]. The carrier rate ranges from as high as 43.6% in Ashkenazi Jewish individuals to 8.5% in East Asians [16], although the number of genes tested and which genes are included are varied among these studies. In our study using 1642 exomes of a Thai population, 39% of Thai individuals carried a PV and an LPV in at least one of 114 genes (113 genes have been recommended by ACMG) [12] plus G6PD, causing G6PD deficiency, which is most common in the Thai population. In this study, 23.8% are carriers of thalassemia and hemoglobinopathies (HBB, HBA1, and HBA2), followed by 7.7% of being a carrier for G6PD deficiency respectively. The carrier rate in our cohort is considered high, similar to the Ashkenazi Jewish group. The most prevalent variants are found in Hb genes and G6PD as aforementioned, are likely the result from the heterozygous advantage in malaria-endemic areas [17] rather than in-group mating as in Ashkenazi Jewish. After excluding thalassemia and hemoglobinopathies, the frequency of the carrier rate in our cohort was still high at 14.7%.

We observed 14 couples (1.7%) who were carriers of PV and LPV in the same gene (S4 Table). Of these couples, seven (0.9%, 7/821) were nonthalassemia carrier couples at risk of having children affected with recessive disorders. Thalassemia carrier screening is already included and available in the screening program of Thai pregnant women; therefore, it is estimated that nearly 1% of the couples could benefit from expanded carrier screening.

The prevalence of common recessive disorders can be estimated in our cohort. In general, this was comparable with the previous estimation of the prevalence rate of common autosomal recessive disorders (thalassemia and G6PD deficiency) [18-24]. This can be implied that the ascertainment and analysis by exclusion of the samples that carry the genetic variants in the disease responsible in their offspring strategy exclude the bias and the frequency rare therefore was comparable to the nondisease or population cohort. This strategy was proven in our previous analysis on the prevalence of the secondary findings. (The majority of the reportable diseases are inherited in an autosomal dominant manner.)

The most common carriers of inherited metabolic disease from our cohort are Gaucher disease caused by mutation in GBA. The result interpretation for this gene is challenging because GBA has a pseudogene. The homology of the GBA pseudogene is highest between exons 8 and 11, which occurs from recombination events [25]. Many clinical exome sequencing laboratories do not analyze or report the GBA variants. The variants commonly identified in our cohort are outside these highly homologous areas and are therefore less likely to be affected in our analysis. Conversely, the prevalence of the GBA variant carrier is likely to be underestimated in our cohort or a similar cohort using exome sequencing.

In our study, carriers of primary hyperoxaluria and Wilson disease PV and LPV were found to be the next most common. Founder variants in East Asian populations (c.2T>C (p.Met1Thr) in AGXT [26-29] and c.2333G>T (p.Arg778Leu) in ATP7B [30-35] were also identified in our cohort. Our study also supported the current newborn screening program in Thailand for Phenylketonuria (PKU) as the estimation of the carrier rates of 1 in 127 individuals comparable with other countries [36-38]; 8–10 children will be born with PKU every year (annual live birth of 600,000).

Hearing loss is one of the major disability problems in childhood, in which hereditary causes both syndromic and nonsyndromic are the main etiologies for these children. We identified that carriers in Pendred syndrome, Usher syndrome, and GJB2 (nonsyndromic hereditary neurosensory hearing loss) were common in Thai cohort similar to previous estimation in other Thai cohorts [39-50].

The carrier of cystic fibrosis in Caucasians are common [51-53], but we identified that only 0.97% of our sample carried PV and LPV in CFTR. As a result of low carrier frequency among Thais, Thais have lower incidence of cystic fibrosis than Caucasians (1/2,000 in Northern European descends vs. 1/400,000) Interestingly, we observed three females (0.18%) bearing 5T (c.1210–12 [5]) of CFTR, which can be found in mild cystic fibrosis but can also result in male infertility due to congenital bilateral absence of the vas deferens [54].

### Limitations

Our study had some limitations. Whole exome sequencing (WES) cannot reliably detect structural variants that are common in α-thalassemia. Spinal muscular atrophy, a common muscular disorder, most commonly caused by a copy number change in the SMN1 gene and modified by the copy numbers of SMN2, is also missed by WES. The genome analysis with structural variant detection will help estimate these common disorders. Additionally, the frequency of variants based on NGS analysis may be underestimated without validation using Sanger sequencing, particularly for complex recombinant alleles of GBA pseudogenes. The other common PVs that will be missed by WES are trinucleotide repeat expansion. PV in FMR1 causing fragile X syndrome, the most common single-gene disorder cause of intellectual disability especially in males, cannot be detected with exome sequencing data. Our sample size has >80% power to detect alleles with frequency of more than 1%. The detection rate for the uncommon variant will require a larger sample size [55].

## Summary

Of the 114 genes, including the 113 ACMG-recommended genes and G6PD, 39% (640/1642) of Thai individuals were found to be carriers. Our study unveiled the landscape of PVs common in a Thai population based on computational filtering and manual curation which will be fundamental for designing a gene list for carrier screening at an individual level or future public health plan.

## Materials and methods

### Study samples

Our study sample included the 1642 exomes of unrelated healthy Thai individuals (811 men and 831 women). Written informed consent was obtained from each individual. This study was approved by the Institutional Review Board of the Faculty of Medicine, Chulalongkorn University (IRB No. 264/62). The data are the part, but not all that were deposited, analyzed, curated in the Thai Reference Exome database [56]. The parents of the patient cohort were enrolled as study samples; therefore we excluded the variants in the genes responsible for patient diagnosis.

### Sequencing and bioinformatics analysis

Genomic DNA was extracted from peripheral blood leukocytes. Exomes were captured using either a TruSeq® Exome Kit (Illumina, San Diego, CA) on the NextSeq 500 system or SureSelect Human All Exons (Agilent Inc., Santa Clara, CA) on the Hiseq 4000 and Novaseq 6000 Systems. The reagents used are according to the manufacturer’s standard protocol. Sequences were aligned to the human reference genome (GRCh37) using the Burrows–Wheeler Aligner package v 0.7.15 [57]. Variant calling was performed using the Genome Analysis Tool Kit (GATK Best Practice V3.7; Broad Institute, CA, USA), which is called by HaplotypeCaller [58]. ANNOVAR was used to annotate genetic variants. The variant interpretation was limited to the 113 ACMG-recommended genes and G6PD (12). We included variants listed as PV and LPV according to VarSome’s ACMG classification [59] with a final manual curation by the authors.

### Statistical analysis

The frequency and 95% confidence interval were calculated using the proportion of carriers in total individuals tested (n/1642) and the Wilson method, respectively.

## Data Availability

All relevant data are within the manuscript and its Supporting Information files

## Acknowledgments

This study was supported by the Health Systems Research Institute (65-040).

## Supporting information

S1 Table. Genetic variants of the 18 parents who were responsible for the presenting symptoms in their children

S2 Table. Carrier frequencies of one or more genetic disorders in 114 genes of 1,642 unrelated Thais

S3 Table. The variant carrier rate (VCR) of each variant

S4 Table. Pathogenic or likely pathogenic variants in the same genes that were harbored by the couples

S5 Table. Characteristics of glucose-6-phosphate dehydrogenase (G6PD) phenotypes in 1,642 unrelated Thais

